# Tattoo ink exposure is associated with lymphoma and skin cancers – a Danish study of twins

**DOI:** 10.1101/2024.07.05.24309993

**Authors:** Signe B. Clemmensen, Jonas Mengel-From, Jaakko Kaprio, Henrik Frederiksen, Jacob vB. Hjelmborg

## Abstract

We aim to study the possible association between tattoo ink exposure and development of certain cancers in the recently established Danish Twin Tattoo Cohort.

Tattoo ink is known to transfer from skin to blood stream and accumulate in regional lymph nodes. We are concerned that tattoo ink induce inflammation at deposit site that may eventually become chronic and increase risk of abnormal cell proliferation, especially skin cancer and lymphoma.

We conducted two designs of study in twins in order to improve confounder control: A cohort study of 2,367 randomly selected twins and a case-control study of 316 twins born in the period 1960-1996. Cancer diagnoses (ICD-10) were retrieved from the Danish Cancer Registry and tattoo ink exposure from the Danish Twin Tattoo survey from 2021. The analysis addressed effects of time-varying exposure.

In the case-control study, individual level analysis resulted in a hazard of skin cancer (of any type except basal cell carcinoma) that was 1.62 times higher among tattooed (95% CI: 1.08-2.41). Twin-matched analysis of 14 twin pairs discordant for tattoo ink exposure and skin cancer show HR=1.33 (95% CI: 0.46-3.84). For skin cancer and lymphoma, increased hazards were found for tattoos larger than the palm of a hand: HR=2.37 (95% CI: 1.11-5.06) and HR=2.73 (95% CI: 1.33-5.60), respectively. In the cohort study design, individual level analysis resulted in a hazard ratio of 3.91 (95% CI: 1.42-10.8) for skin cancer and 2.83 (95% CI: 1.30-6.16) for basal cell carcinoma.

In conclusion, we are very concerned that tattoo ink interacting with surrounding cells may have severe consequences. Further studies are needed beneficial to public health – the sooner the better.

## Introduction

In recent decades, more people have been getting tattooed. The overall proportion of persons with tattoos (i.e. prevalence) is up to 20-25% in some countries – even higher among the younger generations (ref: Thesis Manuscript 3)(1-3). With the increased popularity of tattoos, safety regarding tattooing and exposure to tattoo ink becomes increasingly relevant. Especially the lack of empirical evidence pertaining to carcinogenicity is cause for concern.

Particles from tattoo ink have been found to accumulate in regional lymph nodes (4-6) and they may be transported through the blood stream to other organs (7-9). It is an open question whether they could cause harm in the skin, to the immune system and even to other organs internally.

The most frequently used tattoo ink is black. Black ink typically contains soot products like carbon black which is listed as possibly carcinogenic to humans (mainly based on studies of carbon black inhalation and risk of lung cancer) by the International Agency for Research on Cancer (IARC) (10). Through the incomplete combustion used for carbon black production, polycyclic aromatic hydrocarbons (PAHs) are formed as byproducts, one of the most dangerous being benzo[a]pyrene (BaP) which is classified as carcinogenic to humans by the IARC (11). Another hazardous substance is azo compounds since they may release carcinogenic aromatic amines following exposure to sunlight or laser treatment tattoo removal (12).

The long latency period of cancer and the possible combination of a variety of environmental exposures makes it difficult to describe origin of disease development. The lack of knowledge in this field has caused several researchers along with health-related organizations such as IARC and the European Commission to call for epidemiologic studies to investigate proposed associations between exposure to tattoo ink and risk of certain cancers such as lymphoma and skin cancers (8, 13, 14). To the best of our knowledge, there are only three publications of studies seeking to obtain empirical evidence in regard to this potential link between tattoo ink and risk cancer, more specifically, lymphoma (15, 16), multiple myeloma (15) and basal cell carcinoma (17). The latest study of lymphoma (16) provided evidence suggesting an increased risk among tattooed individuals, while the results from the study of basal cell carcinoma (17) were not statistically significant, though suggestive of an increased odds among tattooed.

We conjecture that tattoo ink induces inflammation at deposit site that may eventually become chronic and increase risk of abnormal cell proliferation, especially skin cancer and lymphoma. We emphasize that this may happen for any type of ink due to foreign body immunologic response. In addition, ink particles with known or suspected carcinogenic properties may gradually increase this risk over time. In this study, we aim to assess the possible relation between tattoo ink exposure and development of certain types of cancer. The cancers considered are those related directly to skin being the original deposit site of tattoo ink along with lymphoma according to the ink deposit conjecture and cancer of the bladder and urinary tract as a deposit site for ink particles transported through the blood stream.

Our recently established Danish Twin Tattoo Cohort provided the opportunity to study described hypothesis with elaborate confounder control. Further, the approach might provide tentative insights to processes governing ink exposures and cancer development expectedly of long duration.

## Materials and methods

The Danish Twin Tattoo Cohort (DTTC) was established in 2021 aiming to collect data for a case-cotwin study and a twin cohort. Subsets of these samples are analyzed in this paper. The DTTC was based on a questionnaire survey about tattoo ink exposure among Danish twins and entitled *Risk factors of certain types of cancer diseases*. The survey was carried out from January to July 2021. Twins were identified through linkage of the nationwide Danish Twin Register (DTR) and the National Cancer Register using unique personal identification numbers. Cancers were categorized according to NORDCAN (18, 19) and thereby comply with Nordic cancer registration. The outcome cancers were a priori chosen to represent our ink deposit conjecture stated in the introduction. Hence, the cancer sites are those for which ink is known or suspected to deposit (4-9). A full description of the cohort compilation can be found in the PhD thesis by Clemmensen (20).

The survey included questions about tattoo status, age at first tattoo, colors, and size (measured in estimated units of the size of the palm of one’s hand). Additionally, there were questions about potential confounders and known cancer risk factors such as smoking, physical exercise, alcohol consumption, and education.

### The case-cotwin study

The twin pairs included in the case-cotwin study were identified as all twin pairs born in Denmark from 1960 to 1996, i.e., all twins had reached age 20 years at end of follow-up (January 1, 2017). Inclusion criteria were: 1) At least one twin had been diagnosed with one (or more) of the following cancers after reaching age 20 years: Hodgkin or non-Hodgkin lymphoma (ICD-10 was used along with ICD-O-3 by NORDCAN when identifying lymphoma incident cases), skin cancer (melanoma ICD-10: C43 and non-melanoma ICD-10: C44 – excluding basal cell carcinoma ICD-10: C44.91) and bladder/urinary tract cancer. 2) At least one twin could be contacted, i.e. was alive, not emigrated, and had not waived contact by researchers. A total of 568 individual twins complied with the first criteria, and 504 individual twins (including 219 complete pairs) further complied with the second criteria and were invited to participate in the survey out of which 316 (56%) responded.

### The twin cohort

The cohort study included 2,459 twin pairs, born 1960-1996, that were selected randomly among all pairs in the DTR where at least one twin could be contacted. A total of 4,532 individual twins were invited and 2,367 (52%) participated (including 673 complete pairs). In the twin cohort, we considered lymphoma, skin cancer, and basal cell carcinoma as outcomes. The lymphoma and skin cancer cases and their cotwins were a subset of the case-cotwin study.

### Statistical analysis

Frequency and percentage or median and interquartile range (IQR) are provided to describe the case-cotwin study along with tattoo size, whether certain colors of ink were used, and smoking habits. Similar information (and more) was reported for the twin cohort previously (ref: Thesis Manuscript 3). Only measured confounders that were known at time of exposure were included. Fewer than five twins had multiple diagnoses; when they were of the same type (e.g. skin cancer), only time to first diagnosis was used, thus disregarding subsequent diagnoses, and when they were of different types (e.g. lymphoma and skin cancer), the individual would appear as a case in both respective samples.

#### Time-to-event analysis

As time to event of cancer is expectedly confounding the studied relationship, the logistic modelling falls short and survival analysis modelling was chosen for the analysis. To assess the hazard ratio of each type of cancer comparing tattoo exposed to non-exposed we applied Cox regression with age as timescale and stratification by sex. Tattoo exposure was included as a time-dependent covariate, meaning that the exposure status of an individual was allowed to change over time, i.e. it changed at time of first tattoo. Thus, an individual acquiring their tattoo after being diagnosed with cancer, only contributed risk time as unexposed. Smoking was added as a risk factor to block potential unmeasured confounders such as lifestyle factors associated with smoking. It was defined as a time-dependent covariate with smoking status changing from non-smoker to smoker at time corresponding to the age of an individual when they started smoking. This will be referred to as never smoker vs ever smoker. We further examined the influence of exposure to certain colors of tattoo ink by comparing the hazard of cancer to those without tattoos while including a term for tattoos without a given color of ink vs no tattoo as a confounder. Similarly, we assessed a dose-response relationship by splitting tattoo exposure into small and large (large defined as larger than the palm of one’s hand).

We aimed to estimate a measure of association for tattoo exposure and adult cancer occurrence (diagnosed from age 20 years). Therefore, the minimum entry age was set to 20 years. Time of censoring was defined as age on January 1, 2017 (end of follow-up on cancer data). In case of multiple diagnoses of the same type, we only considered time to first diagnose, thus disregarding subsequent diagnoses. Scaled Schoenfeld residuals were applied to assess the proportional hazards assumption. Robust sandwich estimation was applied to account for within-pair dependency for twin pairs. As sensitivity analysis, we carried out the same analyses using a frailty model (using gamma-distributed frailties) to assess the influence of different within-pair dependency for monozygotic, same-sex dizygotic, and opposite sex dizygotic twin pairs. Also, as sensitivity analysis we applied inverse probability weight (IPW) adjustment for population representativeness of age and sex in the analysis of the twin cohort as described in (ref: Thesis Manuscript 3) (21).

Finally, we did a matched case-cotwin analysis (also known as a discordant twin design analysis) using a stratified Cox model with twin pair specific baseline hazards to control for unobserved, shared confounding (22). This complements the individual analysis which is more robust towards non-shared confounding (23).

The time-to-event analyses were done using the R packages *survival* (44-46), *timereg* (47, 48), and *mets* (24, 25).

## Results

### Descriptives

Descriptives of the case-cotwin study are displayed in Table 1a. There were 32 lymphoma cases and 34 cotwin controls (from 32 lymphoma discordant twin pairs and 18 incomplete twin pairs). The cases included seven individuals that were tattooed before getting diagnosed with cancer and the median number of years from tattoo to cancer was 8 years (IQR: 4-17 years). Out of the 17 pairs where both twins participated in the survey and provided information on tattoo exposure, there were less than five informative twin pairs (discordant for both lymphoma outcome and tattoo exposure before time of diagnosis in case twin). Among the twin pairs with skin cancer were 119 cases (including 4 concordant twin pairs) and a total of 104 controls (from 67 skin cancer discordant twin pairs and 44 incomplete twin pairs). The cases included 27 individuals that were tattooed before getting diagnosed with cancer and the median number of years from tattoo to cancer was 14 years (IQR: 5-20 years). Out of the 71 pairs where both twins participated in the survey, there were 14 informative twin pairs. Among these were 8 pairs where the case was exposed while the cotwin was unexposed. Among the twin pairs with cancer of the bladder and urinary tract were only 8 cases (all from discordant pairs) and 6 controls. There were not enough informative pairs to provide those counts.

**Table 1a.**
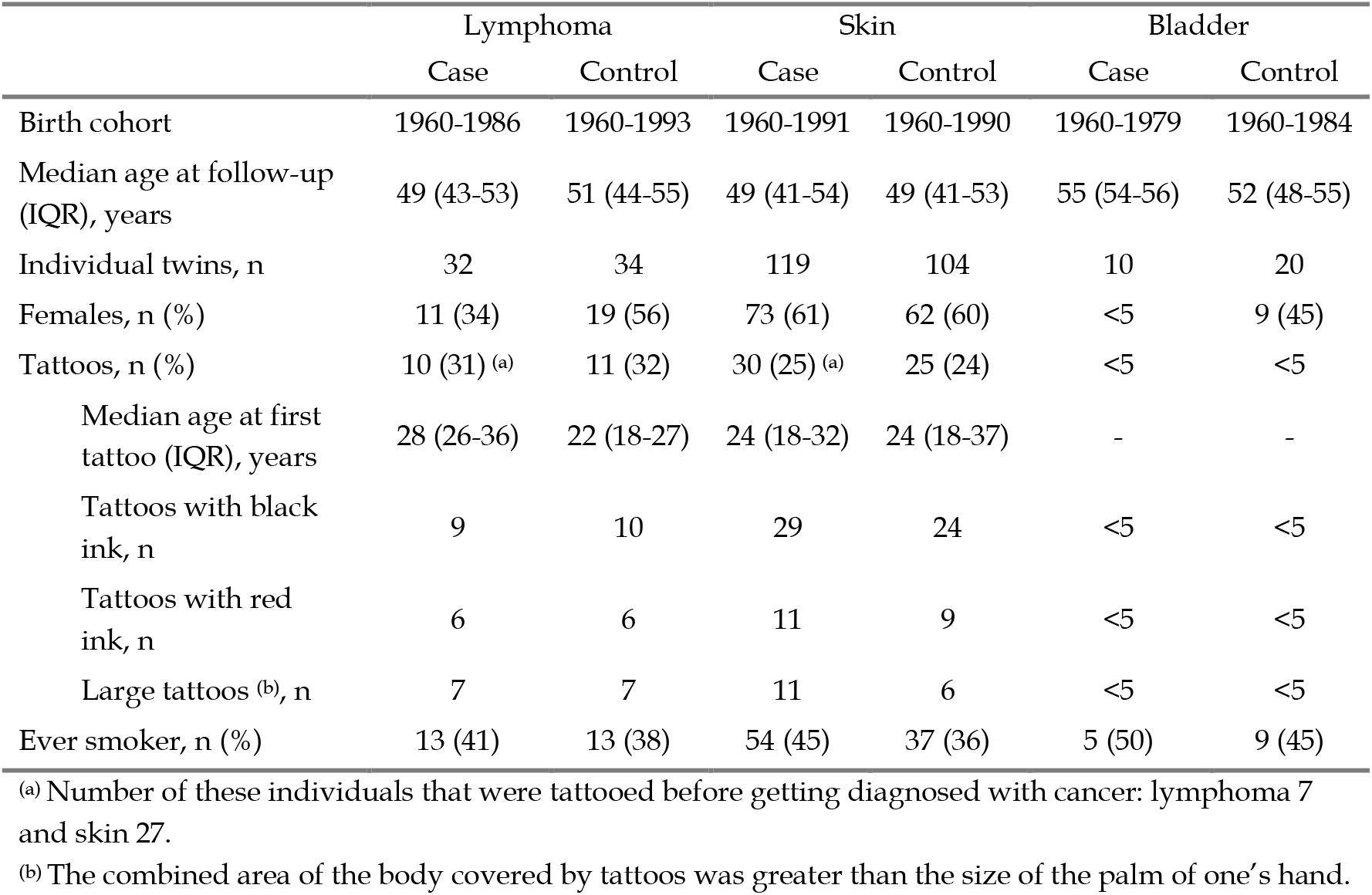
Characteristics of the case-cotwin samples of twin pairs with lymphoma, skin, and bladder cancer. The table includes both twins of complete and incomplete twin-pairs.

|  | Lymphoma |  | Skin |  | Bladder |  |
| --- | --- | --- | --- | --- | --- | --- |
|  | Case | Control | Case | Control | Case | Control |
| Birth cohort | 1960-1986 | 1960-1993 | 1960-1991 | 1960-1990 | 1960-1979 | 1960-1984 |
| Median age at follow-up (IQR), years | 49 (43-53) | 51 (44-55) | 49 (41-54) | 49 (41-53) | 55 (54-56) | 52 (48-55) |
| Individual twins, n | 32 | 34 | 119 | 104 | 10 | 20 |
| Females, n (%) | 11 (34) | 19 (56) | 73 (61) | 62 (60) | <5 | 9 (45) |
| Tattoos, n (%) | 10 (31) <sup>(a)</sup> | 11 (32) | 30 (25) <sup>(a)</sup> | 25 (24) | <5 | <5 |
| Median age at first tattoo (IQR), years | 28 (26-36) | 22 (18-27) | 24 (18-32) | 24 (18-37) | - | - |
| Tattoos with black ink, n | 9 | 10 | 29 | 24 | <5 | <5 |
| Tattoos with red ink, n | 6 | 6 | 11 | 9 | <5 | <5 |
| Large tattoos <sup>(b)</sup>, n | 7 | 7 | 11 | 6 | <5 | <5 |
| Ever smoker, n (%) | 13 (41) | 13 (38) | 54 (45) | 37 (36) | 5 (50) | 9 (45) |

<sup>(a)</sup> Number of these individuals that were tattooed before getting diagnosed with cancer: lymphoma 7 and skin 27.
<sup>(b)</sup> The combined area of the body covered by tattoos was greater than the size of the palm of one's hand.

Descriptives of the twin cohort study are displayed in Table 1b. Among the 2,367 individual twins were 6 cases with lymphoma, 16 with skin cancer (half of them were tattooed with a median number of years from tattoo to diagnosis of 11 years (IQR: 4-20 years)), and 29 with basal cell carcinoma (11 of them were tattooed with a median number of years from tattoo to diagnosis of 14 years (IQR: 7-24 years)). There were 10 individuals among the controls for whom smoking data was missing (<1%). Out of the 673 pairs where both twins participated in the survey and provided information on tattoo exposure, there were less than five informative twin pairs for basal cell carcinoma. The number of informative pairs for lymphoma and skin cancer are not included here, as they are a subset of the case-cotwin samples.

**Table 1b.** Characteristics of the cohort split into cases and controls for lymphoma, skin cancer, and basal cell carcinoma.

|  | Lymphoma |  | Skin |  | Basal Cell Carcinoma |  |
| --- | --- | --- | --- | --- | --- | --- |
|  | Case | Control | Case | Control | Case | Control |
| Birth cohort | 1961-1985 | 1960-1996 | 1960-1991 | 1960-1996 | 1960-1980 | 1960-1996 |
| Median age at follow-up (IQR), years | 50 (44-53) | 42 (30-50) | 50 (46-54) | 42 (30-50) | 52 (49-55) | 41 (29-50) |
| Individual twins, n | 6 | 2,361 | 16 | 2,351 | 29 | 2,338 |
| Females, n (%) | 1-5 | 1,372 (58) | 10 (62) | 1,367 (58) | 19 (66) | 1,358 (58) |
| Tattoo, n (%) | <5 | 664 (28) | 8 (50) <sup>(a)</sup> | 659 (28) | 11 (38) <sup>(a)</sup> | 656 (28) |
| Ever smoker, n (%) | <5 | 858 (36) | 9 (56) | 852 (36) | 8 (28) | 853 (37) |

<sup>(a)</sup> Less than five of these individuals acquired their first tattoo after getting diagnosed with cancer.

In addition to Tables 1a and 1b, information on zygosity and median age at smoking initiation are provided in Supplementary Tables 1a and 1b. Furthermore, Supplementary Tables 2a and 2b display the descriptives of everyone selected for the case-cotwin- and cohort studies that were invited to participate in the survey.

The individuals presented in Tables 1a and 1b are included in the individual level analyses while the informative pairs are considered in the matched analyses. We note that only cases tattooed before getting diagnosed with cancer provides risk time as tattoo exposed in these time-to-event analyses. It will not make sense to perform a two-by-two table analysis of counts in Tables 1a and 1b as the follow-up time is not the same among cases and controls.

### Time-to-event analysis

#### The case-cotwin study

The hazard ratio of lymphoma for large tattoo exposure compared to no tattoo was estimated to 2.73 (95% CI: 1.33-5.60) using an individual level analysis (Table 2). When size was ignored, no evidence of a tattoo effect on hazard of lymphoma could be detected. There was not enough variation to study the effect of tattooing with black ink and no effect of tattooing with red ink was found. It was not possible to fit a model with smoking as covariate or to fit a matched model for lymphoma.

**Table 2.** Hazard ratio of cancer diagnosed from age 20 years among twins born since 1960. Case-cotwin study. Individual level analysis. Stratification by sex has been applied in all models.

|  | Lymphoma |  | Skin |  |
| --- | --- | --- | --- | --- |
|  | HR (95% CI) | p | HR (95% CI) | p |
| Model 1: Tattoo | 1.35 (0.68-2.71) | 0.39 | **1.62** (1.08-2.41) | 0.019 |
| Model 2: Tattoo size |  |  |  |  |
| Small | 0.63 (0.13-2.94) | 0.55 | 1.37 (0.87-2.16) | 0.18 |
| Large | **2.73** (1.33-5.60) | 0.006 | **2.37** (1.11-5.06) | 0.025 |
| Model 3: Red ink |  |  |  |  |
| Yes | 1.36 (0.57-3.29) | 0.49 | 1.44 (0.85-2.45) | 0.18 |
| No | 1.33 (0.36-4.90) | 0.67 | **1.74** (1.02-2.96) | 0.041 |
| Model 4: |  |  |  |  |
| Tattoo | - |  | **1.68** (1.13-2.51) | 0.010 |
| Smoking | - |  | 0.88 (0.62-1.25) | 0.47 |

The hazard ratio of skin cancer for tattoo exposure was estimated to 1.62 (95% CI: 1.08-2.41) in individual level analysis (Table 2). When considering size of tattoo, there was evidence of an effect of large tattoos, HR: 2.37 (95% CI: 1.11-5.06). No effect of exposure to red ink in a tattoo could be detected (HR: 1.44 (95% CI: 0.85-2.45)). There was no evidence of a confounding effect of smoking (ever vs never smoker). The effect of tattoo exposure on hazard of skin cancer in the matched analysis showed HR: 1.33 (95% CI: (0.46-3.84)) and was based on 14 twin pairs discordant for tattoo ink exposure and skin cancer outcome.

It was not possible to estimate the effect of tattooing on hazard of cancer of the bladder and urinary tract.

#### The twin cohort study

The hazard ratio of skin cancer for tattoo exposure was estimated to 3.91 (95% CI: 1.42-10.78) indicating an increased hazard among tattooed individuals (Table 3). Likewise, the hazard ratio of basal cell carcinoma for tattoo exposure was estimated to 2.83 (95% CI: 1.30-6.16). When combining the two cancer types, the hazard ratio was 3.28 (95% CI: 1.76-6.09). There was no evidence of an effect of smoking (ever vs never smoker) when studying skin cancer outcome. For basal cell carcinoma, the effect of tattooing was estimated to HR=3.52 (95% CI: 1.63-7.61) when adjusting for smoking. It was not possible to estimate the effects of tattoo exposure on hazard of lymphoma or to do matched case-cotwin analysis.

**Table 3.**
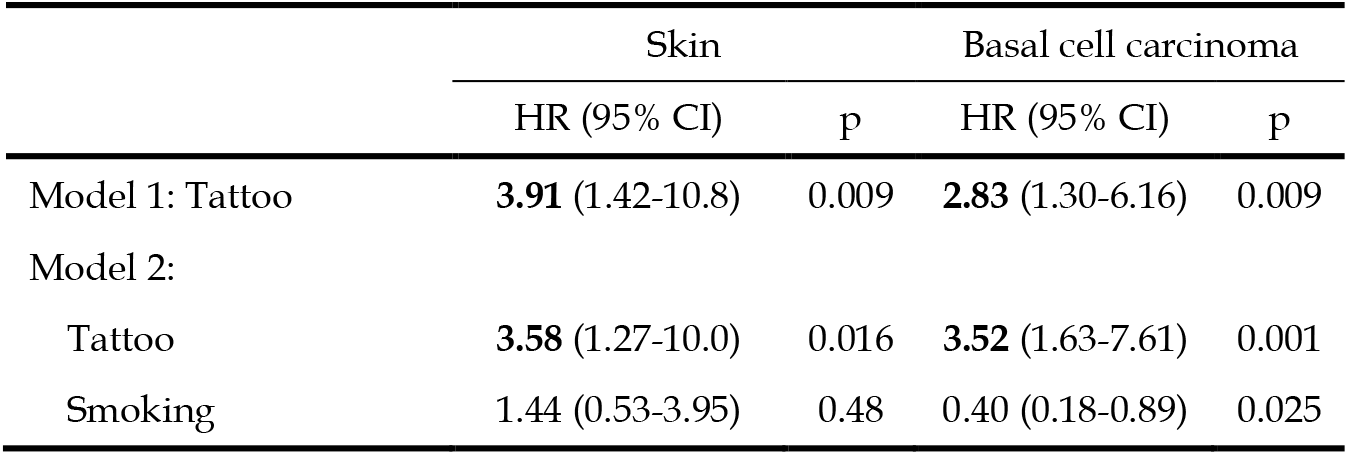
Hazard ratio of cancer diagnosed from age 20 years among twins born since 1960. Cohort study. Individual level analysis. Stratification by sex has been applied in all models.

|  | Skin |  | Basal cell carcinoma |  |
| --- | --- | --- | --- | --- |
|  | HR (95% CI) | p | HR (95% CI) | p |
| Model 1: Tattoo | **3.91** (1.42-10.8) | 0.009 | **2.83** (1.30-6.16) | 0.009 |
| Model 2: |  |  |  |  |
| Tattoo | **3.58** (1.27-10.0) | 0.016 | **3.52** (1.63-7.61) | 0.001 |
| Smoking | 1.44 (0.53-3.95) | 0.48 | 0.40 (0.18-0.89) | 0.025 |

### Sensitivity analysis

Sensitivity analysis indicated no influence of different within-pair dependency for monozygotic, same-sex dizygotic, and opposite sex dizygotic twin pairs. Besides, there were no considerable differences when including IPW adjustment for population representativeness of age and sex in the twin cohort study. Model assumptions (e.g. proportional hazards) were not found violated in any application.

## Discussion

In the case-control study, individual level analysis resulted in a hazard of skin cancer that was 1.62 times higher among tattooed compared to non-tattooed participants (95% CI: 1.08-2.41). The twin-matched analysis of 14 twin pairs discordant for tattoo ink exposure and skin cancer show HR=1.33 (95% CI: 0.46-3.84). For both skin cancer and lymphoma, increased hazards were found for tattoos larger than the palm of a hand: HR=2.37 (95% CI: 1.11-5.06) and HR=2.73 (95% CI: 1.33-5.60), respectively. In the cohort study design, individual level analysis resulted in increased hazards among tattooed for skin cancer, HR=3.91 (95% CI: 1.42-10.8), and basal cell carcinoma, HR=2.83 (95% CI: 1.30-6.16).

A strength of our study is the application of two designs of studies with complementary advantages allowing for extended confounder control. The twin sample provides a highly valid and representative control group e.g. in terms of age, sex, upbringing, and genetic similarity. Additional merits of our study are: i) The application of time-to-event analysis using age as time scale to precisely specify risk time according to age of being tattooed or non-tattooed at time of diagnosis and thus avoiding immortal time bias and bias from incomplete follow-up. In general, considering a time-varying covariate as constant, typically leads to underestimation of exposure effect. ii) Using inverse probability weights for age and sex representativeness for better confounder adjustment (compared to standard covariate adjustment).

Our sample consists of cancer survivors participating in the survey. Those passing away, e.g. due to severe cancer, will not be represented. This group consists of less than 10% of the eligible twins, including emigrated and those waivered contact by researchers. We suspect limited survivorship bias.

There are several different types of tattoos. The intended focus of this study was “classic” decorative tattoos. However, since the survey did not specify type of tattoo to the participants, individuals with permanent make-up (PMU) and medical tattoos may also have responded as being tattooed. In hindsight, there should have been a question distinguishing these types of tattoos as they generally differ in both size and the type of ink used.

When assessing the influence of smoking, the time-varying covariate was defined only from age when the participants started smoking and assumed one never stopped smoking. Here only crude smoking information was included, and we did not include e.g. number of pack years or, in a matched analysis, by considering the total number of years as a smoker until time of diagnose in the case twin.

The case-control sample of twin pairs with cancer of the bladder and urinary tract expectedly did not contain enough cases to study the effect of tattoo ink exposure. It is possibly too early to study this association since bladder cancer mainly occurs at high age, that is, among individuals from generations where tattoo prevalence is currently low as demonstrated in (ref: Thesis manuscript 3).

Our study was initiated on the basis of the suspicion that ink deposits will interact with surrounding tissue causing increased cell proliferation and thereby increase cancer risk. We term this *the ink deposit conjecture*. The mechanism involves an immunologic response and is recognized for instance in breast implant-associated anaplastic large cell lymphoma (BIA-ALCL), a rare type of T-cell lymphoma (26). We stress that this pathway does not necessarily involve particular ink agents, however, if carcinogenic compounds are present the pathway is expectedly different but still leads to increased cancer risk. Consequently, the preventive effects of the recent European restrictions (27) intended to limit exposure to a long list of known or suspected carcinogenic compounds may be lower than first anticipated. Our findings are consistent with the conjecture and with reported findings such as squamous cell carcinoma, benign tumors, lymphoid conditions, and rare cases of malignant neoplasms occurring within the area of a tattoo (28-33).

To the best of our knowledge, there are only three publications in the field as of June 2024. The first is a study from 2020 about cosmetic tattooing and early onset basal cell carcinoma in New Hampshire (17). The study sample was based on a matched case-control design, but only exposed individuals (156 tattooed cases and 213 tattooed controls) were included in the analysis. They compared odds of being tattooed within the “anatomical region” of the basal cell carcinoma (as opposed to being tattooed at a different site) to odds of being tattooed within randomly assigned “reference sites” and found an odds ratio of 1.8 (95% CI: 1.0-3.2) hinting towards association. The second study is a Canadian study from 2020 (15) and considered two population-based case-control studies holding 1,518 participants (737 cases) from a study of non-Hodgkin lymphoma and 742 (373 cases) from a study of multiple myeloma. Using logistic regression modelling, they found no association with tattoo exposure. The third study, a Swedish population-based case-control study of lymphoma from 2024 (16) included 1,398 cases and 4,193 controls identified through incidence density sampling. Their main result of increased risk of lymphoma among tattooed is borderline significant. Through conditional logistic regression (i.e. matched analysis) using basic and extended confounder adjustment, they estimate incidence rate ratios (IRR) of lymphoma of 1.21 (95% CI: 0.99-1.48) and 1.24 (95% CI: 1.02-1.50), respectively, suggesting increased risk among tattooed individuals. A strength of the study is, that they obtain very similar IRR’s in both unadjusted and adjusted, matched and individual level analysis. The authors comment on estimates relating to dose-response relationships and influence of exposure time, but the results presented provide no evidence to support this discussion.

The more recently born individuals in the Nordic countries do not have markedly higher age-specific incidence rates of non-Hodgkin lymphoma and non-melanoma skin cancer, thus not supporting a major role of tattoos on cancer incidence in the population (18, 19). It may be that the proportion of cases accounted for by the increase in incidence for tattoos is too small to be detected in the overall variation of cancer incidence. For skin cancer, it may be balanced out by e.g. increasing use of methods and behaviors to decrease sun exposure.

Having a tattoo, especially among adolescents, has been suggested as an indicator of risky lifestyle highly associated with e.g. smoking (ref: thesis manuscript 3) and alcohol consumption (34) – both risk factors of certain cancer types. Hence, evidence of an association between tattoo ink exposure and occurrence of cancer may be confounded by other health related lifestyle factors. We intend to exploit the remainder of the information gathered in the survey in the future – both regarding lifestyle factors, but also the tattoo details, e.g. is it possible to point at one particular color of tattoo ink? Additionally, there is potential for updated follow-up and extension of the project in the future. For instance, through linkage with national disease registers, one could study association between tattoo ink exposure and incidence of other diseases of the immune system.

The present study was restricted to individuals born since 1960 to avoid bias from birth cohort differences. However, the Danish Twin Tattoo Cohort holds information dating back since the time of the opening of the Danish Cancer Register in 1943. Thus, it would be possible to extend the follow-up period, but alternative means of analysis would be required. It is difficult to model association between tattoo exposure and cancer incidence over such a long period because i) the popularity of tattooing has increased markedly in recent decades (ref: thesis manuscript 3) and ii) cancer diagnostic procedures have improved markedly over the years. Additionally, the incidence of both tattooing and cancer varies with age. That is, two timescales are in play: individual age and calendar time. Through Poisson modelling it is possible to estimate a hazard ratio of cancer by tattoo exposure while incorporating both timescales. Also, to enable matched case-cotwin modelling, (expectedly the strongest possible design for studying exposure-outcome association), larger cohorts are needed. There is potential to expand the Danish Twin Tattoo Cohort to include the other Nordic countries through their respective twin registers.

With the increasing popularity of getting tattooed, it is necessary to question the safety of laser tattoo removal where pigments are broken into smaller fragments that leave the site of the tattoo. The question is: where do the pigment fragments end up? Decreasing particle size often allows for greater migration potential (35). This is also an issue in relation to decomposition of ink particles induced by sun radiation (12). Besides, with ink particles travelling through the blood, could tattooing during pregnancy or the following period of breast feeding be injurious to health of the offspring? Further, on the more speculative side, we suggest research into ink compositions that can be dissolved or investigations into medical treatments that can remove some of the persistent chemicals from the body, especially the lymphatic system.

In conclusion, we have studied tattoo ink exposure and occurrence of certain cancers among Danish twins using two designs: a twin cohort and a case-cotwin study. We are very concerned that tattoo ink has severe public health consequences since tattooing is very abundant among the younger generation.

## Data Availability

All data produced in the present study are available upon reasonable request to the Danish Twin Registry.

## Statements and declarations

## Author contributions

All authors contributed to the study conception and design. Material preparation, data collection and analysis were performed by Signe B Clemmensen, and Jacob vB Hjelmborg. The first draft of the manuscript was written by Signe B Clemmensen and all authors commented on previous versions of the manuscript. All authors read and approved the final manuscript.

## Acknowledgements

The authors are thankful to The Danish Twin Registry for aiding in the collection and managing the data from the Danish Twin Tattoo Cohort. We also acknowledge the contribution of all of the participants of the survey.

## Data Availability Statement

Restrictions apply to the availability of these data. Requests to access data need to be made through the Danish Twin Registry.

### Ethics approval

Ethical approval was waived by the Regional Committees on Health Research and Ethics for Southern Denmark. The appropriate register authorities have given permission for the record linkages.

### Informed consent statement

Informed consent was obtained from all individual participants included in the study.

## Supplementary materials

**Supplementary Table 1a.**
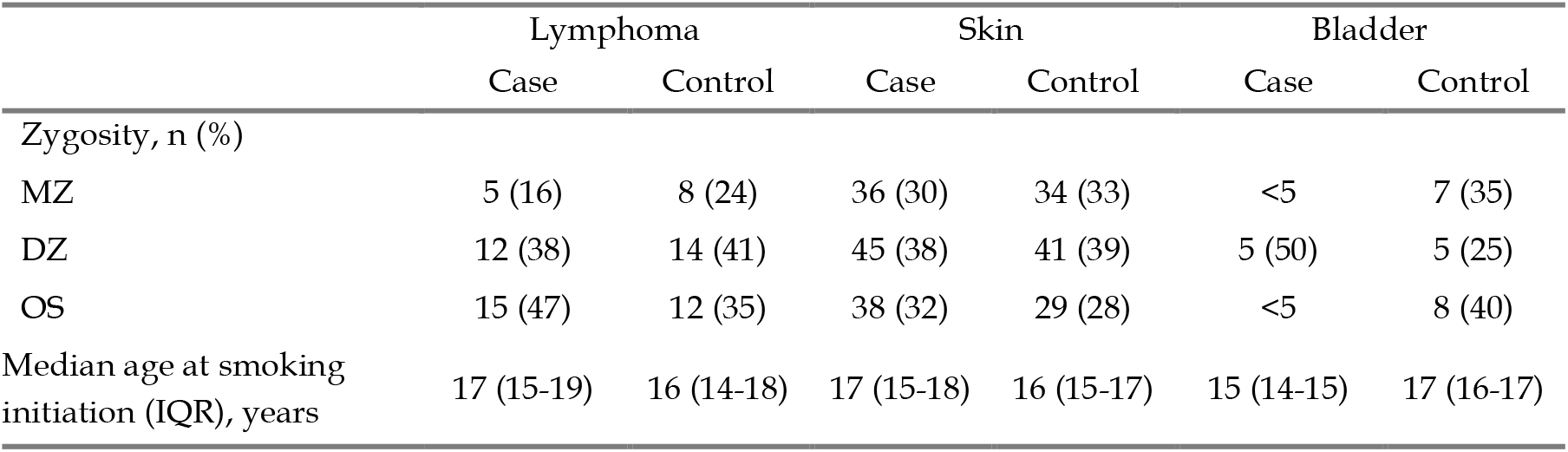
Characteristics of the case-cotwin samples of twin pairs with lymphoma, skin, and bladder cancer. Additional to Table 1a.

**Supplementary Table 1b.** Characteristics of the cohort split into cases and controls for lymphoma, skin cancer, and basal cell carcinoma. Additional to Table 1b.

|  | Lymphoma |  | Skin |  | Basal Cell Carcinoma |  |
| --- | --- | --- | --- | --- | --- | --- |
|  | Case | Control | Case | Control | Case | Control |
| Zygosity, n (%) |  |  |  |  |  |  |
| MZ | <5 | 636 (27) | <5 | 632 (27) | 11 (38) | 626 (27) |
| DZ | <5 | 866 (37) | 9 (56) | 861 (37) | 11 (38) | 859 (37) |
| OS | <5 | 859 (36) | <5 | 858 (36) | 7 (24) | 853 (36) |
| Median age at smoking initiation (IQR), years | - | 16 (14-18) | 17 (17-25) | 16 (14-18) | 16 (14-17) | 16 (14-18) |

**Supplementary Table 2a.** Characteristics of the case-cotwin samples of twin pairs invited to participate in the survey where at least one twin in a pair was diagnosed with lymphoma, skin, or bladder cancer.

|  | Lymphoma |  | Skin |  | Bladder |  |
| --- | --- | --- | --- | --- | --- | --- |
|  | Case | Control | Case | Control | Case | Control |
| Birth cohort | 1960-1996 | 1960-1996 | 1960-1994 | 1960-1994 | 1960-1984 | 1960-1984 |
| Median age at follow-up (IQR), years | 49 (43-53) | 49 (43-54) | 48 (41-53) | 48 (40-53) | 53 (49-55) | 53 (48-55) |
| Individual twins, n | 48 | 57 | 182 | 175 | 24 | 24 |
| Females, n (%) | 17 (35) | 31 (54) | 109 (60) | 100 (57) | <5 | 10 (42) |
| Zygosity, n (%) |  |  |  |  |  |  |
| MZ | 8 (17) | 12 (21) | 51 (28) | 53 (30) | 7 (29) | 9 (38) |
| DZ | 19 (40) | 21 (37) | 73 (40) | 66 (38) | 11 (46) | 7 (29) |
| OS | 21 (44) | 24 (42) | 58 (32) | 56 (32) | 6 (25) | 8 (35) |
| Survey participation, n (%) | 32 (67) | 34 (60) | 119 (65) | 104 (59) | 10 (42) | 20 (83) |

**Supplementary Table 2b.** Characteristics of the cohort for all twins invited to participate in the survey split into twins with and without lymphoma, skin cancer or basal cell carcinoma.

|  | Lymphoma |  | Skin |  | Basal cell carcinoma |  |
| --- | --- | --- | --- | --- | --- | --- |
|  | Case | Control | Case | Control | Case | Control |
| Birth cohort | 1961-1985 | 1960-1996 | 1960-1991 | 1960-1996 | 1960-1993 | 1960-1996 |
| Median age at follow-up (IQR), years | 51 (44-52) | 40 (28-49) | 49 (46-52) | 40 (28-49) | 50 (46-53) | 40 (27-49) |
| Individual twins, n | 10 | 4,522 | 27 | 4,505 | 45 | 4,487 |
| Females, n (%) | 6 (60) | 2,299 (51) | 15 (56) | 2,290 (51) | 26 (58) | 2,279 (51) |
| Zygosity, n (%) |  |  |  |  |  |  |
| MZ | 1-5 | 1,156 (26) | 9 (33) | 1,150 (26) | 17 (38) | 1,142 (25) |
| DZ | 5 (50) | 1,666 (37) | 13 (48) | 1,658 (37) | 17 (38) | 1,654 (37) |
| OS | 1-5 | 1,700 (38) | 5 (19) | 1,697 (38) | 11 (24) | 1,691 (38) |
| Survey participation, n (%) | 6 (60) | 2,362 (52) | 16 (59) | 2,352 (52) | 29 (64) | 2,339 (52) |

